# Genetic susceptibility versus fibrosis progression in North Indian MASLD: distinct roles of APOC3 and PNPLA3 in a candidate gene study

**DOI:** 10.64898/2026.02.25.26347059

**Authors:** Nandini Tomar, Sangeeta Choudhury, Anil Arora, Praveen Sharma, Rishikant Vaibhav, Rizwana Hasan, Shayesta Jan, Rajwinder Kaur, Tarun Rajput, Medha Sravanthi Lomada, Sandhya Kiran Pemmasani, Ashish Kumar

## Abstract

**Background and Aim:** MASLD affects 30–38% of Indian adults, yet the contribution of genetic risk variants to disease susceptibility and fibrosis progression remains poorly characterised. We investigated the association of 12 candidate SNPs with MASLD susceptibility and fibrosis severity in North Indian patients, benchmarking allele frequencies against IndiGenomes and global populations.

**Methods:** Sixty-nine MASLD patients (75.4% male; median BMI 29.8 kg/m²) from a tertiary care liver clinic in New Delhi were genotyped for 12 SNPs using Illumina custom BeadChip array and Sanger sequencing. Patients were stratified by liver stiffness measurement (LSM): significant fibrosis (≥8 kPa, n=38) versus no significant fibrosis (<8 kPa, n=31). Allele frequencies were compared with IndiGenomes (∼1,020 Indian individuals) and 1000 Genomes populations.

**Results:** *PNPLA3* rs738409 G allele was the strongest within-cohort predictor of significant fibrosis (allelic OR 2.89, 95% CI 1.35–6.19, P=0.006; dominant model OR 3.94, P=0.008), with carriers demonstrating higher LSM (median 15.6 vs. 7.5 kPa, P=0.005). *SAMM50* rs3761472 (OR 2.12, P=0.065) and *FTO* rs9939609 (OR 2.08, P=0.089) showed non-significant trends. In the population-level comparison, *APOC3* rs2854116 T allele was the only variant significantly enriched after Bonferroni correction (64.0% vs. 47.9%; OR 1.93, 95% CI 1.35–2.77, P<0.001), followed by *PNPLA3* (33.3% vs. 24.1%, OR 1.57, P=0.019) and *SAMM50* (31.2% vs. 22.6%, OR 1.55, P=0.028). Notably, *APOC3* showed no association with fibrosis (OR 0.96, P=1.000), suggesting a role in susceptibility rather than progression. All SNPs were in Hardy-Weinberg equilibrium.

**Conclusions:** This study reveals a dissociation between genetic determinants of MASLD susceptibility and fibrosis progression in North Indian patients. *APOC3* rs2854116 predisposes to MASLD at the population level, while *PNPLA3* rs738409 drives fibrosis severity within established disease, underscoring the need for ancestry-specific genetic risk stratification.

**Graphical Abstract:** 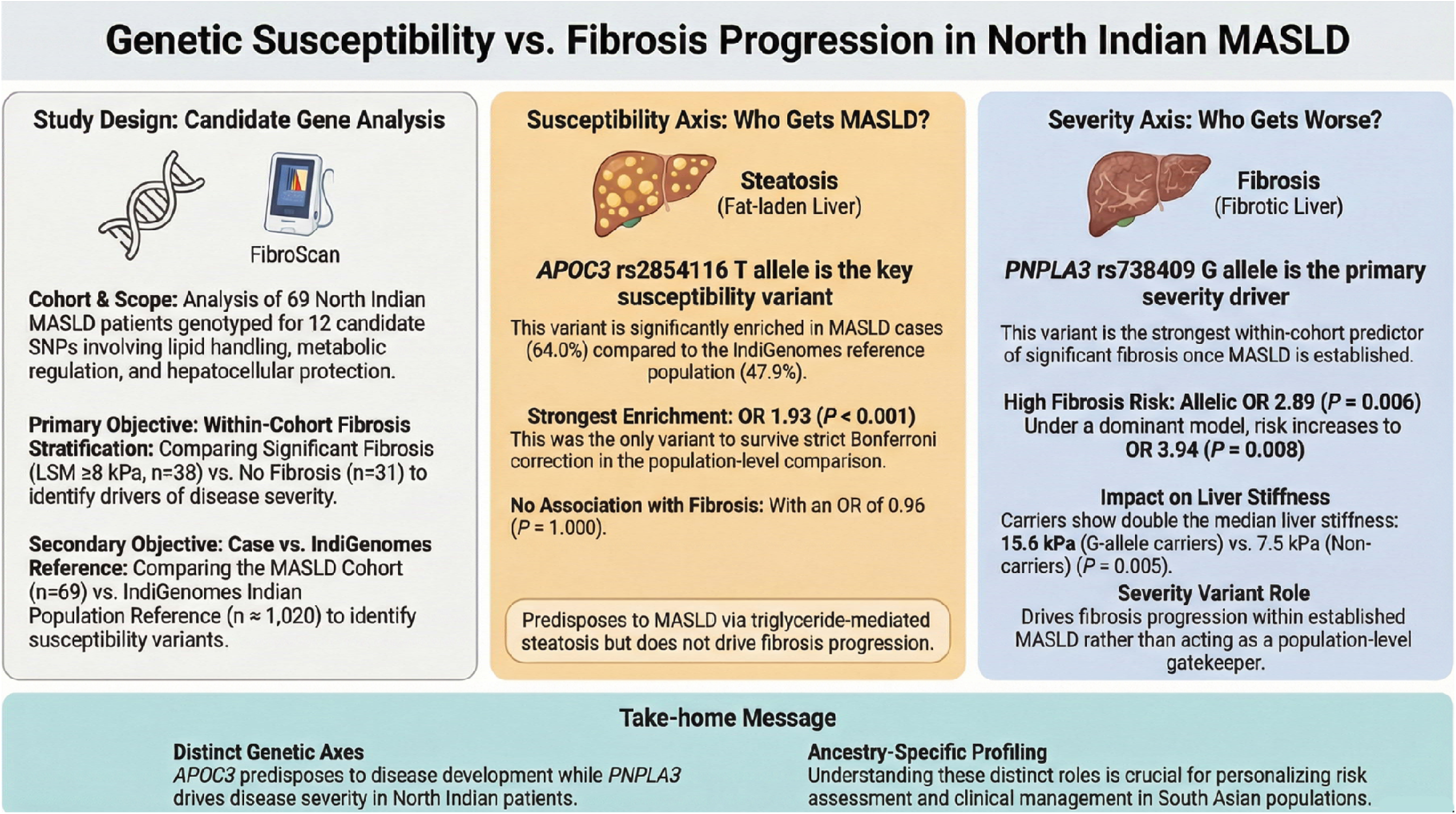

## INTRODUCTION

Metabolic dysfunction–associated steatotic liver disease (MASLD), formerly termed non-alcoholic fatty liver disease (NAFLD), is the leading chronic liver disorder worldwide, affecting approximately 30–40% of the adult population globally^1–7^. The disease encompasses a spectrum ranging from simple hepatic steatosis to metabolic dysfunction–associated steatohepatitis (MASH), progressive fibrosis, cirrhosis, and hepatocellular carcinoma^8,9^. Among South Asians, MASLD is particularly prevalent and presents at lower body mass indices than in Western cohorts, reflecting the ‘thin-fat’ phenotype characterised by central adiposity and heightened insulin resistance^10–12^. In India, adult MASLD prevalence ranges from 30 to 38%, with rapid urbanisation, dietary transitions, and sedentary lifestyles driving an escalating disease burden^7,13,14^.

MASLD is a polygenic condition in which host genetic variation interacts with metabolic and environmental factors to govern hepatic lipid accumulation, inflammation, and fibrogenesis. Over the past decade, genome-wide association studies and candidate gene analyses have identified several robust susceptibility loci ^15,16^. PNPLA3 rs738409 (p.Ile148Met), the most widely replicated variant, impairs hepatocyte triglyceride lipolysis and confers a 1.5–3-fold risk of steatosis and fibrosis across diverse populations^17–19^. TM6SF2 rs58542926 (p.Glu167Lys) disrupts hepatic very-low-density lipoprotein secretion^20–22^, while MBOAT7 variants alter phospholipid remodelling and synergise with PNPLA3 in promoting fibrosis progression^23,24^. The protective HSD17B13 rs72613567 splice variant reduces hepatocyte lipotoxicity^25,26^. Additional loci including GCKR rs1260326, FTO rs9939609, TCF7L2 rs7903146, LEPR rs1137101, MC4R rs17782313, SAMM50 rs3761472, ATP2A1 rs3888190, and APOC3 rs2854116 have shown variable associations with steatosis, obesity, diabetes, or dyslipidaemia in context-dependent patterns^27–31^.

Critically, the vast majority of these genetic discoveries derive from European and East Asian populations, where allele frequencies, haplotype structures, and gene–environment interactions differ substantially from those observed in South Asians^32,33^. Genetic architecture varies across ancestries due to population-specific minor allele frequencies, linkage disequilibrium patterns, and metabolic contexts, rendering risk models developed in Western populations potentially unreliable for Indian patients. The IndiGenomes project provides whole-genome sequencing data from 1,029 self-declared healthy individuals representing diverse Indian ethnic groups and geographic regions, offering an ancestry-relevant reference against which disease-associated allele enrichment can be evaluated^34^.

The association between MASLD and genetic variants in Indian populations remains incompletely characterised. A limited number of Indian studies have examined variants such as PNPLA3 and HSD17B13, with inconsistent results across cohorts^35–41^. Whether the broader panel of established MASLD-associated variants is enriched in Indian patients, and whether these variants predict fibrosis severity or merely disease susceptibility, has not been systematically addressed.

We therefore undertook a candidate gene study in 69 North Indian patients with clinically and FibroScan-characterised MASLD, genotyping a 12-SNP panel that spans lipid handling (PNPLA3, TM6SF2, MBOAT7, APOC3), metabolic regulation (GCKR, FTO, TCF7L2, LEPR, MC4R), hepatocellular protection (HSD17B13, SAMM50), and calcium homeostasis (ATP2A1). The study pursued two objectives. The primary objective was to identify genotype–phenotype associations within the cohort by comparing patients with significant fibrosis (LSM ≥8 kPa) against those without, thereby identifying variants that drive disease severity once MASLD is established. The secondary objective was to compare allele frequencies between the MASLD cohort and the IndiGenomes reference population^34^, identifying variants enriched in MASLD patients relative to the general Indian population and thus potentially involved in disease susceptibility. This dual-objective design addresses two distinct biological questions—who gets worse, and who gets the disease—using a single genotyped cohort.

## METHODS

### Study Design and Setting

This prospective, single-centre genetic association study was conducted at the Fatty Liver and Obesity Clinic of the Institute of Liver, Gastroenterology & Pancreatico-Biliary Sciences (ILGPS), Sir Ganga Ram Hospital, New Delhi in association with Department of Biotechnology and Research, Sir Ganga Ram Hospital, New Delhi, India. The clinic, operational since January 2024, follows a multidisciplinary model involving hepatologists, dietitians, and exercise physiologists, providing integrated care for MASLD and related metabolic disorders through lifestyle interventions, medical therapy, and ongoing research.

The study protocol was approved by the Institutional Ethics Committee of Sir Ganga Ram Hospital, New Delhi (Approval No. EC/07/25/2735). All participants provided written informed consent prior to saliva collection and genotyping. The study was conducted in accordance with the Declaration of Helsinki (2013 revision) and applicable Indian regulatory guidelines. Patient data were de-identified prior to analysis.

### Study Participants

Consecutive adult patients (aged ≥18 years) attending the Fatty Liver and Obesity Clinic between August 2025 and Feb 2026 who were willing to undergo genetic testing were screened for eligibility. MASLD was diagnosed on the basis of ultrasound evidence of hepatic steatosis in the presence of at least one cardiometabolic risk factor, in accordance with the recent multi-society Delphi consensus definition^3^. Cardiometabolic risk factors included overweight or obesity (BMI ≥23 kg/m² per Asian criteria), type 2 diabetes mellitus (T2DM), hypertension, and dyslipidaemia.

Patients were excluded if they had significant alcohol consumption (>14 standard drinks per week for women or >21 per week for men), Metabolic Dysfunction-Associated Alcoholic Liver Disease (MetALD), alcoholic liver disease, viral hepatitis (hepatitis B or C), autoimmune hepatitis, hereditary liver diseases (Wilson disease, haemochromatosis), primary biliary cholangitis, drug-induced liver injury, biliary obstruction, severe end-organ damage, human immunodeficiency virus infection, pregnancy, or lactation. Patients with incomplete clinical records or genotyping failure were also excluded.

A total of 69 patients met the inclusion criteria and formed the final study cohort.

### Clinical and Anthropometric Assessment

A detailed clinical history was obtained for each participant, including demographic data, socioeconomic profile, smoking and alcohol intake history, physical activity patterns, and family history of T2DM, obesity, hypertension, liver disease, and coronary artery disease. Height, weight, waist circumference, and hip circumference were measured using standard protocols, and body mass index (BMI) was calculated as weight (kg) divided by height (m) squared.

### Liver Stiffness and Steatosis Assessment

Transient elastography (FibroScan, Echosens, Paris, France) was performed for all participants to assess liver stiffness measurement (LSM, in kPa) as a surrogate for hepatic fibrosis and controlled attenuation parameter (CAP, in dB/m) as a measure of hepatic steatosis. Only examinations with at least 10 valid measurements and an interquartile range to median ratio ≤0.30 were considered reliable. Significant fibrosis was defined as LSM ≥8 kPa, and advanced fibrosis or cirrhosis as LSM ≥12 kPa. Significant steatosis was defined as CAP ≥248 dB/m, in accordance with published thresholds for Indian populations.

### Definition of Comorbidities

Comorbid conditions were defined using established clinical criteria. Obesity was defined as BMI ≥25 kg/m² (Asian criteria) or waist circumference ≥90 cm in males and ≥80 cm in females. T2DM was defined as a prior diagnosis on treatment, or fasting blood glucose ≥126 mg/dL, or HbA1c ≥6.5%. Hypertension was defined as a prior diagnosis on treatment, or blood pressure ≥130/85 mmHg. Dyslipidaemia was defined as serum triglycerides ≥150 mg/dL, or HDL-C ≤40 mg/dL in males and ≤50 mg/dL in females, or LDL-C ≥160 mg/dL, or current use of lipid-lowering therapy.

### Selection of Candidate Genetic Variants

Twelve single nucleotide polymorphisms (SNPs) across established MASLD-related genes were selected through a structured review of genome-wide association studies, candidate gene analyses, and meta-analyses (PubMed, Scopus, Web of Science; up to March 2024). Variants were chosen on the basis of reproducibility across populations, biological plausibility, and prior associations with hepatic steatosis, fibrosis progression, or related metabolic traits. The panel included PNPLA3 rs738409 (I148M), HSD17B13 rs72613567, TM6SF2 rs58542926, MBOAT7 rs626283, GCKR rs1260326, FTO rs9939609, TCF7L2 rs7903146, LEPR rs1137101, ATP2A1 rs3888190, MC4R rs17782313, APOC3 rs2854116, and SAMM50 rs3761472. These variants span pathways involved in hepatic lipid handling (PNPLA3, TM6SF2, MBOAT7, APOC3), metabolic regulation (GCKR, FTO, TCF7L2, LEPR, MC4R), proteostasis (HSD17B13, SAMM50), and calcium homeostasis (ATP2A1). The complete list of SNPs with flanking sequences and supporting references is provided in Supplementary Table S1. Of note, the canonical MBOAT7 variant associated with MASLD in European GWAS is rs641738; the variant genotyped in this study (rs626283) is located in the same gene region and has been independently associated with hepatic steatosis, although its linkage disequilibrium with rs641738 in South Asian populations requires further characterisation.

### DNA Extraction and Genotyping

Saliva samples were collected from all participants using the MapmyGenome at-home DNA collection kit (Cat # CY-93050R-TS) following the manufacturer’s protocol. Genomic DNA was extracted using a standard salting-out method, quantified by NanoDrop spectrophotometry (ThermoFisher, USA) (A260/A280 ratio >1.8), and normalised to 50 ng/μL.

Genotyping for 11 of the 12 SNPs was performed using the Illumina Infinium iSelect HTS Custom Genotyping BeadChip-24 (Catalog ID: WG-405-1014), a custom microarray comprising 10,133 probes targeting 8,768 unique variants. The assay was carried out following the manufacturer’s protocol: briefly, genomic DNA underwent whole-genome amplification, enzymatic fragmentation, precipitation, and resuspension prior to hybridisation onto the beadchip. After single-base extension and staining, imaging was performed using the Illumina high-throughput scanner. Raw intensity data (.idat files) were processed using the Illumina GenomeStudio Genotyping Module v2.0 (Illumina Inc., USA) for genotype clustering, allele calling, and quality assessment.

For HSD17B13 rs72613567, which was not represented on the custom array, polymerase chain reaction followed by Sanger sequencing was performed. Primers were designed using the NCBI Primer-BLAST tool (Forward: 5′-AATGCCTCGCCACCATTTTG-3′; Reverse: 5′-CCCCAGGGATGGAGAGTTTC-3′), yielding an amplicon of 816 bp. PCR amplification used Takara Emerald PCR Master Mix (Cat # RR320A) under optimised cycling conditions. Amplified products were verified by agarose gel electrophoresis, purified, and sequenced using BigDye Terminator chemistry (Applied Biosystems) on an ABI 3730 DNA Analyzer (Cat # A41046). Genotypes were assigned from sequencing chromatograms based on observed nucleotide peaks.

Samples with genotyping call rates below 95% on the Illumina array were excluded from analysis.

### Reference Population

Allele and genotype frequencies for a population-level reference were obtained from the IndiGenomes database, which comprises whole-genome sequencing data from 1,029 healthy, unrelated individuals of diverse Indian ancestry. IndiGenomes provides ancestry-resolved variant frequencies that serve as a valuable benchmark for case-enrichment analyses in Indian populations. Allele and genotype counts for each of the 12 SNPs were extracted from the IndiGenomes browser (build GRCh38). Sample sizes varied slightly across SNPs depending on data availability in the database (range: n ≈ 1,014 to 1,026).

It should be noted that IndiGenomes participants were not individually phenotyped for MASLD or its components. Given that the prevalence of MASLD in Indian adults is estimated at 30–38%, a proportion of reference individuals may harbour subclinical hepatic steatosis. This potential misclassification would attenuate observed case-reference allele frequency differences and bias odds ratios toward the null. Furthermore, individual-level data on age, sex, BMI, and metabolic parameters were not available from IndiGenomes, precluding matched or adjusted comparisons. Additionally, the IndiGenomes cohort represents diverse Indian ethnicities, whereas our study cohort comprises North Indian subjects; residual population stratification cannot be entirely excluded. These limitations are acknowledged in the interpretation of findings from the case-reference analysis.

### Statistical Analysis

All statistical analyses were performed using Python (version 3.13) with SciPy (scipy.stats module) for Fisher’s exact test, Mann-Whitney U test, chi-squared test, and Shapiro-Wilk normality testing. Odds ratios and 95% confidence intervals were calculated using Woolf’s method. Data management and preliminary computations were carried out in Microsoft Excel. Figures were generated using Matplotlib (version 3.x) in Python.

For the primary objective (genotype-phenotype associations within the cohort), patients were stratified by fibrosis status — significant fibrosis (LSM ≥8 kPa) versus non-significant fibrosis (LSM <8 kPa). Genotype and allele frequency distributions for each SNP were compared between these groups using Pearson’s chi-squared test or Fisher’s exact test, as appropriate based on expected cell counts. Odds ratios with 95% confidence intervals were calculated for allelic, dominant (carriers of at least one risk allele versus homozygous reference), and recessive (homozygous risk versus others) genetic models. The prevalence of cardiometabolic comorbidities (obesity, T2DM, hypertension, dyslipidaemia) was compared across genotype groups using chi-squared or Fisher’s exact test. Continuous biochemical and anthropometric variables were compared by genotype using independent-samples t-test or Mann-Whitney U test, depending on the distribution of data assessed by the Shapiro-Wilk test. Given the exploratory sample size (n = 69), multivariable logistic regression was not performed, as the number of events per variable would fall below conventional thresholds for model stability. Associations between genotype and fibrosis status were therefore assessed using Fisher’s exact test across allelic, dominant, and recessive genetic models, which provide assumption-free inference appropriate for small samples.

For the secondary objective (case-reference allele frequency comparison), allele and genotype frequencies in the study cohort were compared with IndiGenomes reference frequencies using the chi-squared test. Odds ratios with 95% confidence intervals were calculated for allelic associations. Risk allele frequencies were additionally compared with global population estimates from the 1000 Genomes Project and the Genome Aggregation Database (gnomAD) across continental ancestry groups (African, admixed American, East Asian, South Asian, and European) to contextualise population-level differences.

Hardy-Weinberg equilibrium (HWE) was assessed for all 12 SNPs in both the study cohort and IndiGenomes controls using exact tests. Any SNP deviating from HWE at P<0.001 in the reference population was flagged for potential genotyping error.

To account for multiple comparisons across 12 SNPs, the Bonferroni-corrected significance threshold was set at P<0.004 (0.05/12) for the primary allelic analyses. Nominal significance (P<0.05) is also reported for completeness and to guide hypothesis generation, with explicit labelling of results that do not withstand correction. A post-hoc power calculation was conducted using G*Power 3.1. For the within-cohort fibrosis analysis (38 versus 31 patients), the study had 80% power to detect an allelic odds ratio of approximately 2.5 at α = 0.05 for a variant with minor allele frequency of 30%. For the case–reference analysis (69 cases versus 1,020 controls), the study had 80% power to detect an allelic odds ratio of approximately 1.7 at the Bonferroni-corrected threshold of P < 0.004 for a variant with minor allele frequency of 25%. Power limitations for smaller effect sizes are acknowledged in the interpretation of non-significant results.

Continuous data are presented as mean ± standard deviation (SD) for normally distributed variables or median with interquartile range (IQR) for skewed distributions. Categorical data are presented as counts and percentages. A two-tailed P<0.05 was considered nominally significant unless otherwise specified.

## RESULTS

### Baseline characteristics

The study cohort comprised 69 patients with MASLD, of whom 52 (75.4%) were male. The mean age was 47.7 ± 9.3 years and the median BMI was 29.8 kg/m² (IQR 27.8–33.1), with 62 patients (89.9%) meeting the Asian obesity criterion of BMI ≥25 kg/m². Comorbidity prevalence was substantial: type 2 diabetes mellitus in 22 (31.9%), hypertension in 28 (40.6%), and dyslipidaemia in 46 (66.7%). Thirty-eight patients (55.1%) had significant fibrosis (LSM ≥8 kPa), including 26 (37.7%) with advanced fibrosis (LSM ≥12 kPa). Sixty patients (87.0%) had significant hepatic steatosis on CAP (≥248 dB/m).

Patients with significant fibrosis were heavier (median weight 88.0 vs. 78.2 kg, P = 0.005), had higher BMI (31.0 vs. 28.5 kg/m², P = 0.011), greater waist circumference (113.5 vs. 103.0 cm, P = 0.003), and a higher waist-to-height ratio (0.67 vs. 0.61, P = 0.023) than those without significant fibrosis. Type 2 diabetes was significantly more prevalent in the fibrosis group (44.7% vs. 16.1%, P = 0.023). CAP values were paradoxically lower in patients with significant fibrosis (mean 277.5 vs. 306.5 dB/m, P = 0.008), consistent with the well-described phenomenon of steatosis regression accompanying fibrosis progression in advanced MASLD. The complete baseline characteristics stratified by fibrosis status are presented in Table 1.

**Table 1:**
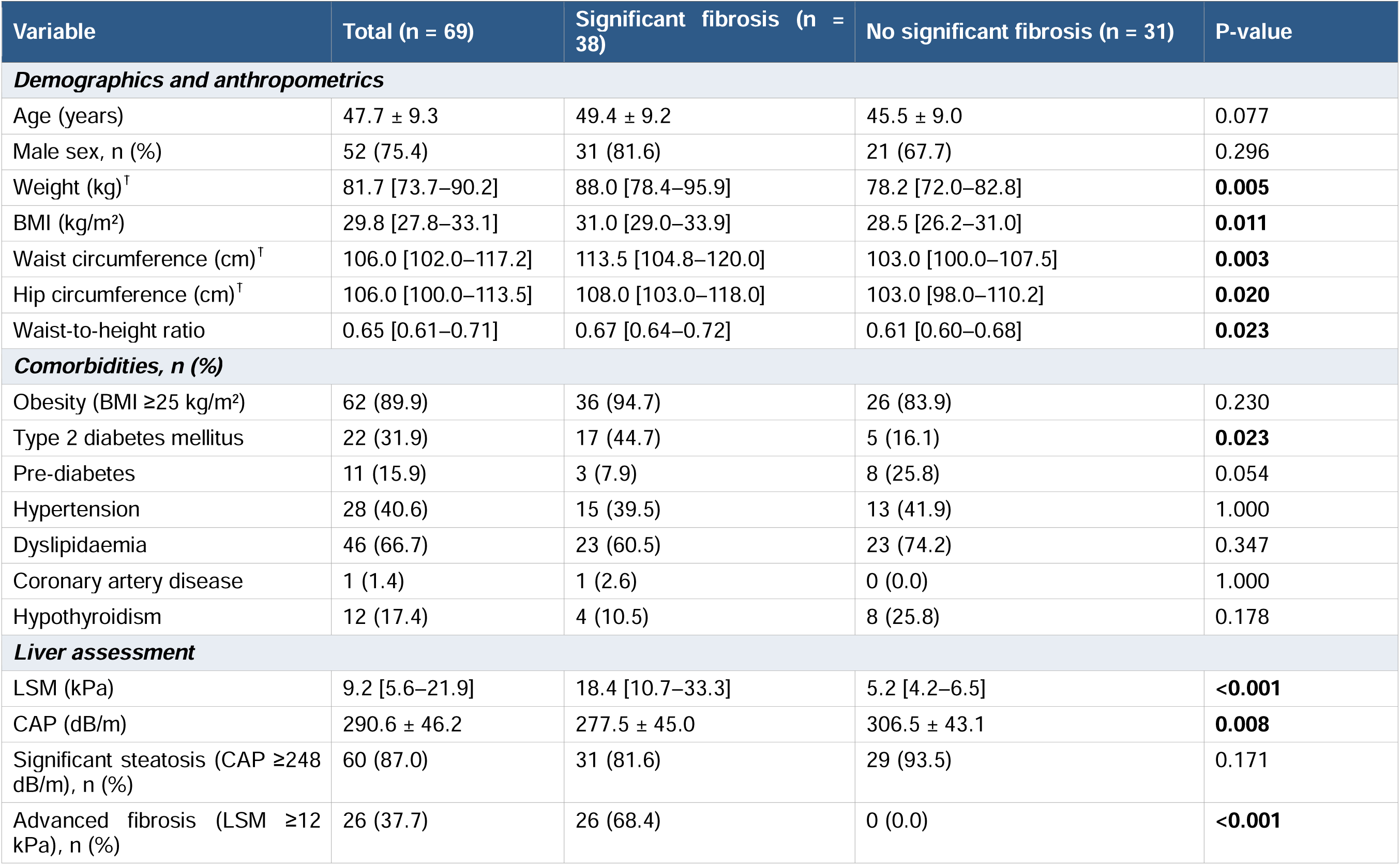
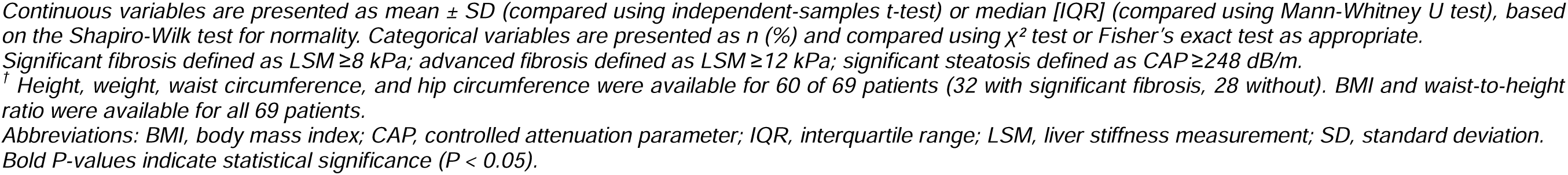
Baseline demographic, anthropometric, and clinical characteristics of the study cohort (n = 69), stratified by liver fibrosis status.

| Variable | Total (n = 69) | Significant fibrosis (n = 38) | No significant fibrosis (n = 31) | P-value |
| --- | --- | --- | --- | --- |
| **Demographics and anthropometrics** |  |  |  |  |
| Age (years) | 47.7 ± 9.3 | 49.4 ± 9.2 | 45.5 ± 9.0 | 0.077 |
| Male sex, n (%) | 52 (75.4) | 31 (81.6) | 21 (67.7) | 0.296 |
| Weight (kg)<sup>†</sup> | 81.7 [73.7–90.2] | 88.0 [78.4–95.9] | 78.2 [72.0–82.8] | **0.005** |
| BMI (kg/m<sup>2</sup>) | 29.8 [27.8–33.1] | 31.0 [29.0–33.9] | 28.5 [26.2–31.0] | **0.011** |
| Waist circumference (cm)<sup>†</sup> | 106.0 [102.0–117.2] | 113.5 [104.8–120.0] | 103.0 [100.0–107.5] | **0.003** |
| Hip circumference (cm)<sup>†</sup> | 106.0 [100.0–113.5] | 108.0 [103.0–118.0] | 103.0 [98.0–110.2] | **0.020** |
| Waist-to-height ratio | 0.65 [0.61–0.71] | 0.67 [0.64–0.72] | 0.61 [0.60–0.68] | **0.023** |
| **Comorbidities, n (%)** |  |  |  |  |
| Obesity (BMI ≥25 kg/m<sup>2</sup>) | 62 (89.9) | 36 (94.7) | 26 (83.9) | 0.230 |
| Type 2 diabetes mellitus | 22 (31.9) | 17 (44.7) | 5 (16.1) | **0.023** |
| Pre-diabetes | 11 (15.9) | 3 (7.9) | 8 (25.8) | 0.054 |
| Hypertension | 28 (40.6) | 15 (39.5) | 13 (41.9) | 1.000 |
| Dyslipidaemia | 46 (66.7) | 23 (60.5) | 23 (74.2) | 0.347 |
| Coronary artery disease | 1 (1.4) | 1 (2.6) | 0 (0.0) | 1.000 |
| Hypothyroidism | 12 (17.4) | 4 (10.5) | 8 (25.8) | 0.178 |
| **Liver assessment** |  |  |  |  |
| LSM (kPa) | 9.2 [5.6–21.9] | 18.4 [10.7–33.3] | 5.2 [4.2–6.5] | **<0.001** |
| CAP (dB/m) | 290.6 ± 46.2 | 277.5 ± 45.0 | 306.5 ± 43.1 | **0.008** |
| Significant steatosis (CAP ≥248 dB/m), n (%) | 60 (87.0) | 31 (81.6) | 29 (93.5) | 0.171 |
| Advanced fibrosis (LSM ≥12 kPa), n (%) | 26 (37.7) | 26 (68.4) | 0 (0.0) | **<0.001** |

*Continuous variables are presented as mean ± SD (compared using independent-samples t-test) or median [IQR] (compared using Mann-Whitney U test), based on the Shapiro-Wilk test for normality. Categorical variables are presented as n (%) and compared using $\chi^2$ test or Fisher's exact test as appropriate.*
*Significant fibrosis defined as LSM ≥ 8 kPa; advanced fibrosis defined as LSM ≥ 12 kPa; significant steatosis defined as CAP ≥ 248 dB/m.*
*<sup>†</sup> Height, weight, waist circumference, and hip circumference were available for 60 of 69 patients (32 with significant fibrosis, 28 without). BMI and waist-to-height ratio were available for all 69 patients.*
*Abbreviations: BMI, body mass index; CAP, controlled attenuation parameter; IQR, interquartile range; LSM, liver stiffness measurement; SD, standard deviation.*
*Bold P-values indicate statistical significance ( $P < 0.05$ ).*

### Genotype distribution and Hardy-Weinberg equilibrium

Genotype data were available for all 69 patients for 11 of 12 SNPs; one patient had missing data for *APOC3* rs2854116 (n = 68 for this locus). All 12 SNPs were in Hardy-Weinberg equilibrium in the combined cohort (all HWE P > 0.05), indicating no genotyping artefacts or systematic bias. Genotype distributions are detailed in Table 2.

**Table 2:**
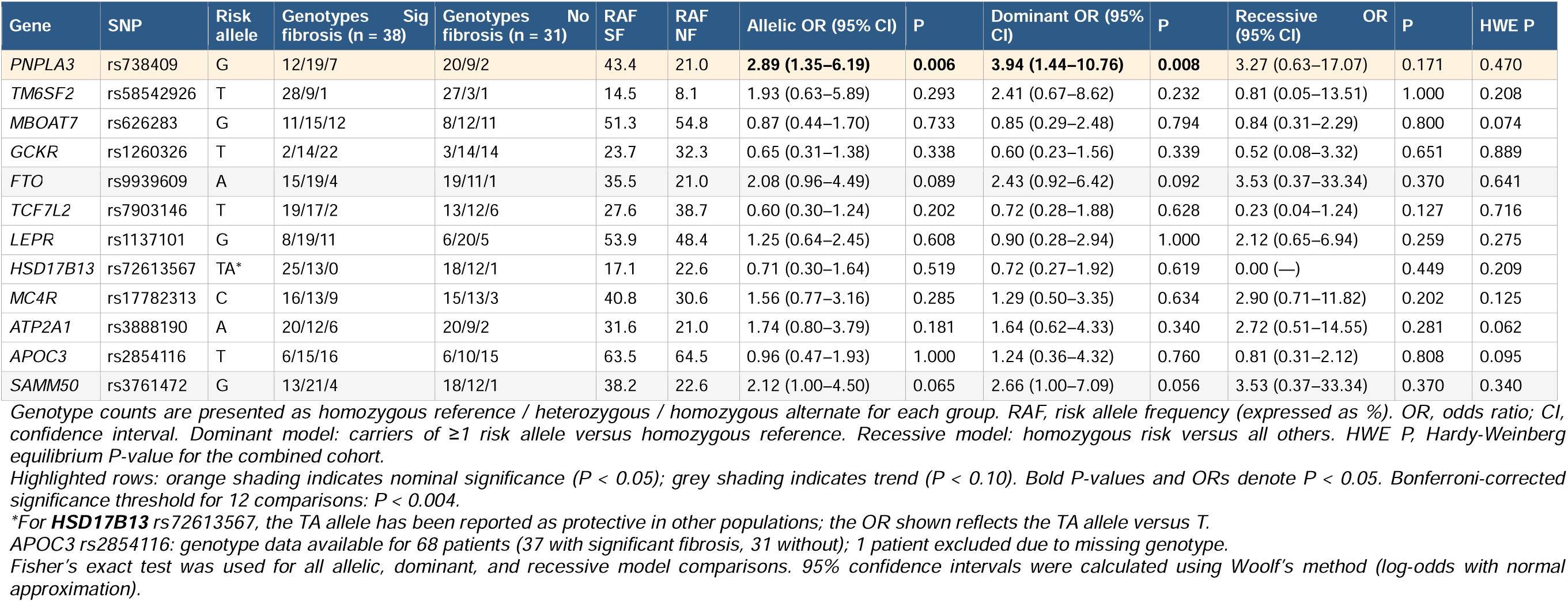
Association of candidate genetic variants with significant liver fibrosis (LSM ≥8 kPa) in patients with MASLD (n = 69).

### Genetic associations with significant fibrosis (primary objective)

Among the 12 candidate SNPs tested for association with significant fibrosis (LSM ≥8 kPa), PNPLA3 rs738409 emerged as the strongest signal. The G (risk) allele frequency was 43.4% in the significant fibrosis group compared with 21.0% in the no-fibrosis group, yielding an allelic odds ratio of 2.89 (95% CI 1.35–6.19, P = 0.006). Under the dominant model, carriers of at least one G allele (CG or GG genotype) had nearly fourfold higher odds of significant fibrosis than CC homozygotes (OR 3.94, 95% CI 1.44–10.76, P = 0.008). While this association did not reach the Bonferroni-corrected threshold of P < 0.004, it was robust at the nominal level and consistent across genetic models.

Two additional variants showed borderline trends toward fibrosis association. SAMM50 rs3761472 G allele was more frequent in the fibrosis group (38.2% vs. 22.6%, allelic OR 2.12, 95% CI 1.00–4.50, P = 0.065), and under the dominant model carriers trended toward significance (OR 2.66, P = 0.056). FTO rs9939609 A allele also trended higher in the fibrosis group (35.5% vs. 21.0%, OR 2.08, 95% CI 0.96–4.49, P = 0.089). The remaining nine SNPs, including APOC3 rs2854116 (OR 0.96, P = 1.000), TM6SF2 rs58542926 (OR 1.93, P = 0.293), and TCF7L2 rs7903146 (OR 0.60, P = 0.202), showed no significant association with fibrosis status. The complete results across all genetic models are presented in Table 2 and the forest plot of allelic odds ratios in Figure 1.

**Figure 1:**
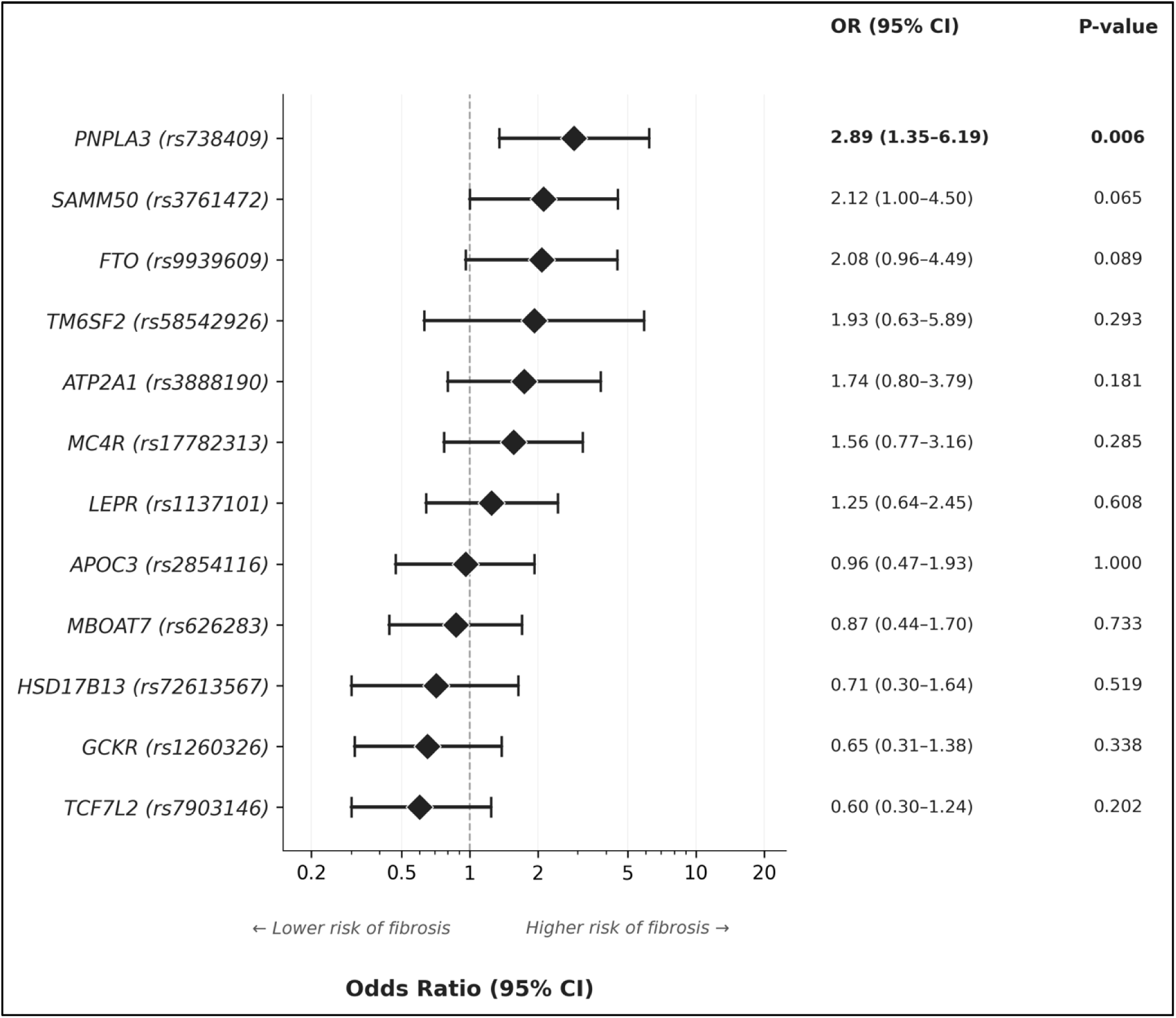
Forest plot of allelic associations with significant liver fibrosis (LSM ≥8 kPa) for 12 candidate SNPs in North Indian patients with MASLD (n = 69). Diamond markers represent allelic odds ratios; horizontal lines indicate 95% confidence intervals. The dashed vertical line denotes the null value (OR = 1). *PNPLA3* rs738409 G allele was the only variant with a nominally significant association with significant fibrosis (OR 2.89, 95% CI 1.35–6.19, P = 0.006). *SAMM50* rs3761472 (OR 2.12, P = 0.065) and *FTO* rs9939609 (OR 2.08, P = 0.089) showed borderline trends. No variant reached the Bonferroni-corrected threshold of P < 0.004. Odds ratios were calculated using Fisher’s exact test with Woolf’s method for 95% confidence intervals. SNPs are ordered by descending odds ratio.

### Genotype–phenotype associations with comorbidities and liver parameters

To explore the broader phenotypic correlates of risk genotypes, we examined associations between carrier status (dominant model) and metabolic comorbidities, BMI, LSM, and CAP for the five SNPs with at least one nominally significant or borderline finding (Table 3).

**Table 3:** Genotype–phenotype associations: comorbidities and liver parameters stratified by carrier status for selected candidate variants (dominant model).

| Variable | Carriers | Non-carriers | OR (95% CI) | P |
| --- | --- | --- | --- | --- |
| **PNPLA3 rs738409** | **G carriers (CG+GG) (n = 37)** | **CC (n = 32)** | **OR (95% CI)** | **P** |
| Type 2 diabetes, n (%) | 15 (40.5) | 7 (21.9) | 2.44 (0.83–7.22) | 0.124 |
| Hypertension, n (%) | 18 (48.6) | 10 (31.2) | 2.08 (0.77–5.66) | 0.219 |
| Dyslipidaemia, n (%) | 26 (70.3) | 20 (62.5) | 1.42 (0.51–3.94) | 0.610 |
| Hypothyroidism, n (%) | 3 (8.1) | 9 (28.1) | 0.23 (0.05–0.94) | 0.053 |
| Sig fibrosis (LSM ≥8 kPa), n (%) | 26 (70.3) | 12 (37.5) | 3.94 (1.44–10.76) | **0.008** |
| LSM (kPa), median [IQR] | 15.6 [7.0–30.5] | 7.5 [5.1–9.7] | **P = 0.005** |  |
| CAP (dB/m), median [IQR] | 278.0 [255.0–309.0] | 305.0 [281.8–337.5] | **P = 0.025** |  |
| BMI (kg/m<sup>2</sup>), median [IQR] | 30.5 [27.9–33.1] | 29.2 [27.8–32.9] | P = 0.504 |  |
| **SAMM50 rs3761472** | **G carriers (AG+GG) (n = 38)** | **AA (n = 31)** | **OR (95% CI)** | **P** |
| Type 2 diabetes, n (%) | 14 (36.8) | 8 (25.8) | 1.68 (0.58–4.85) | 0.437 |
| Hypertension, n (%) | 14 (36.8) | 14 (45.2) | 0.71 (0.27–1.88) | 0.623 |
| Dyslipidaemia, n (%) | 25 (65.8) | 21 (67.7) | 0.92 (0.33–2.55) | 1.000 |
| Obesity (BMI ≥25 kg/m<sup>2</sup>), n (%) | 37 (97.4) | 25 (80.6) | 8.88 (1.04–75.78) | **0.040** |
| Sig fibrosis (LSM ≥8 kPa), n (%) | 25 (65.8) | 13 (41.9) | 2.66 (1.00–7.09) | 0.056 |
| LSM (kPa), median [IQR] | 12.8 [6.3–28.2] | 7.6 [5.0–10.9] | P = 0.067 |  |
| CAP (dB/m), median [IQR] | 284.5 [260.8–314.8] | 301.0 [275.5–318.0] | P = 0.359 |  |
| BMI (kg/m<sup>2</sup>), median [IQR] | 30.5 [28.2–32.7] | 29.3 [27.0–33.4] | P = 0.503 |  |
| **FTO rs9939609** | **A carriers (AT+AA) (n = 35)** | **TT (n = 34)** | **OR (95% CI)** | **P** |
| Type 2 diabetes, n (%) | 11 (31.4) | 11 (32.4) | 0.96 (0.34–2.68) | 1.000 |
| Hypertension, n (%) | 19 (54.3) | 9 (26.5) | 3.30 (1.17–9.29) | **0.027** |
| Dyslipidaemia, n (%) | 22 (62.9) | 24 (70.6) | 0.71 (0.26–1.93) | 0.611 |
| Sig fibrosis (LSM ≥8 kPa), n (%) | 23 (65.7) | 15 (44.1) | 2.43 (0.92–6.42) | 0.092 |
| LSM (kPa), median [IQR] | 11.8 [5.6–27.6] | 7.8 [5.7–14.6] | P = 0.101 |  |
| CAP (dB/m), median [IQR] | 299.0 [269.0–316.5] | 287.5 [263.8–315.0] | P = 0.540 |  |
| BMI (kg/m<sup>2</sup>), median [IQR] | 30.6 [28.5–33.9] | 29.1 [27.2–31.5] | P = 0.114 |  |
| **TM6SF2 rs58542926** | **T carriers (CT+TT) (n = 14)** | **CC (n = 55)** | **OR (95% CI)** | **P** |
| Type 2 diabetes, n (%) | 7 (50.0) | 15 (27.3) | 2.67 (0.80–8.90) | 0.120 |
| Hypertension, n (%) | 1 (7.1) | 27 (49.1) | 0.08 (0.01–0.66) | **0.005** |
| Dyslipidaemia, n (%) | 8 (57.1) | 38 (69.1) | 0.60 (0.18–2.00) | 0.527 |
| Sig fibrosis (LSM ≥8 kPa), n (%) | 10 (71.4) | 28 (50.9) | 2.41 (0.67–8.62) | 0.232 |
| LSM (kPa), median [IQR] | 12.8 [8.2–28.8] | 8.0 [5.1–20.5] | P = 0.095 |  |
| CAP (dB/m), median [IQR] | 283.0 [266.2–303.0] | 299.0 [262.0–320.0] | P = 0.371 |  |
| BMI (kg/m<sup>2</sup>), median [IQR] | 29.5 [27.4–30.9] | 30.0 [27.9–33.4] | P = 0.408 |  |
| **APOC3 rs2854116 †** | **T carriers (TC+TT) (n = 56)** | **CC (n = 12)** | **OR (95% CI)** | **P** |
| Type 2 diabetes, n (%) | 17 (30.4) | 5 (41.7) | 0.61 (0.17–2.26) | 0.505 |
| Hypertension, n (%) | 23 (41.1) | 5 (41.7) | 0.98 (0.27–3.50) | 1.000 |
| Dyslipidaemia, n (%) | 38 (67.9) | 7 (58.3) | 1.51 (0.42–5.39) | 0.522 |
| Sig fibrosis (LSM ≥8 kPa), n (%) | 31 (55.4) | 6 (50.0) | 1.24 (0.36–4.32) | 0.760 |
| LSM (kPa), median [IQR] | 9.2 [5.5–22.2] | 8.1 [5.9–14.6] | P = 0.624 |  |
| CAP (dB/m), median [IQR] | 292.5 [265.2–319.5] | 303.5 [267.8–314.2] | P = 0.797 |  |
| BMI (kg/m<sup>2</sup>), median [IQR] | 30.0 [28.4–33.5] | 27.6 [26.3–32.3] | P = 0.084 |  |

Dominant genetic model: carriers of ≥ 1 risk allele versus homozygous reference genotype. Categorical variables compared using Fisher's exact test; continuous variables compared using Mann-Whitney U test. OR, odds ratio; CI, confidence interval; IQR, interquartile range; LSM, liver stiffness measurement; CAP, controlled attenuation parameter.
† APOC3 rs2854116: n = 68 (1 missing genotype); carriers n = 56, non-carriers n = 12.

PNPLA3 rs738409 G carriers demonstrated the most consistent genotype–phenotype pattern. Beyond their association with significant fibrosis (70.3% vs. 37.5%, P = 0.008), carriers had significantly higher median LSM (15.6 vs. 7.5 kPa, P = 0.005) and paradoxically lower median CAP (278.0 vs. 305.0 dB/m, P = 0.025), reinforcing the concept that steatosis may regress as fibrosis advances in PNPLA3-associated MASLD. A borderline inverse association with hypothyroidism was also noted (8.1% vs. 28.1% in non-carriers, P = 0.053), though this should be interpreted cautiously given the small numbers.

SAMM50 rs3761472 G carriers showed a striking association with obesity (97.4% vs. 80.6% in non-carriers, OR 8.88, P = 0.040) and borderline trends toward higher fibrosis prevalence (65.8% vs. 41.9%, P = 0.056) and elevated LSM (median 12.8 vs. 7.6 kPa, P = 0.067). FTO rs9939609 A carriers had significantly higher hypertension prevalence (54.3% vs. 26.5%, OR 3.30, P = 0.027), consistent with the established role of FTO in metabolic syndrome. An unexpected finding was an inverse association between TM6SF2 rs58542926 T carrier status and hypertension (7.1% vs. 49.1%, P = 0.005), though only 14 patients carried the T allele, warranting cautious interpretation. APOC3 rs2854116 showed no significant within-cohort associations with any comorbidity, liver parameter, or fibrosis measure, further supporting the interpretation that this variant influences disease susceptibility at the population level rather than disease severity.

### Case–IndiGenomes allele frequency comparison (secondary objective)

Comparison of risk allele frequencies between the MASLD cohort and the IndiGenomes reference population revealed three variants with significant or near-significant enrichment in cases (Table 4, Figure 2). APOC3 rs2854116 was the only variant to survive Bonferroni correction: the T allele was present at 64.0% in cases versus 47.9% in IndiGenomes (OR 1.93, 95% CI 1.35–2.77, P < 0.001). This represents a substantial enrichment and suggests that the APOC3 promoter variant, which upregulates apolipoprotein C-III expression and promotes hypertriglyceridaemia, confers genetic susceptibility to MASLD in the Indian population.

**Figure 2:**
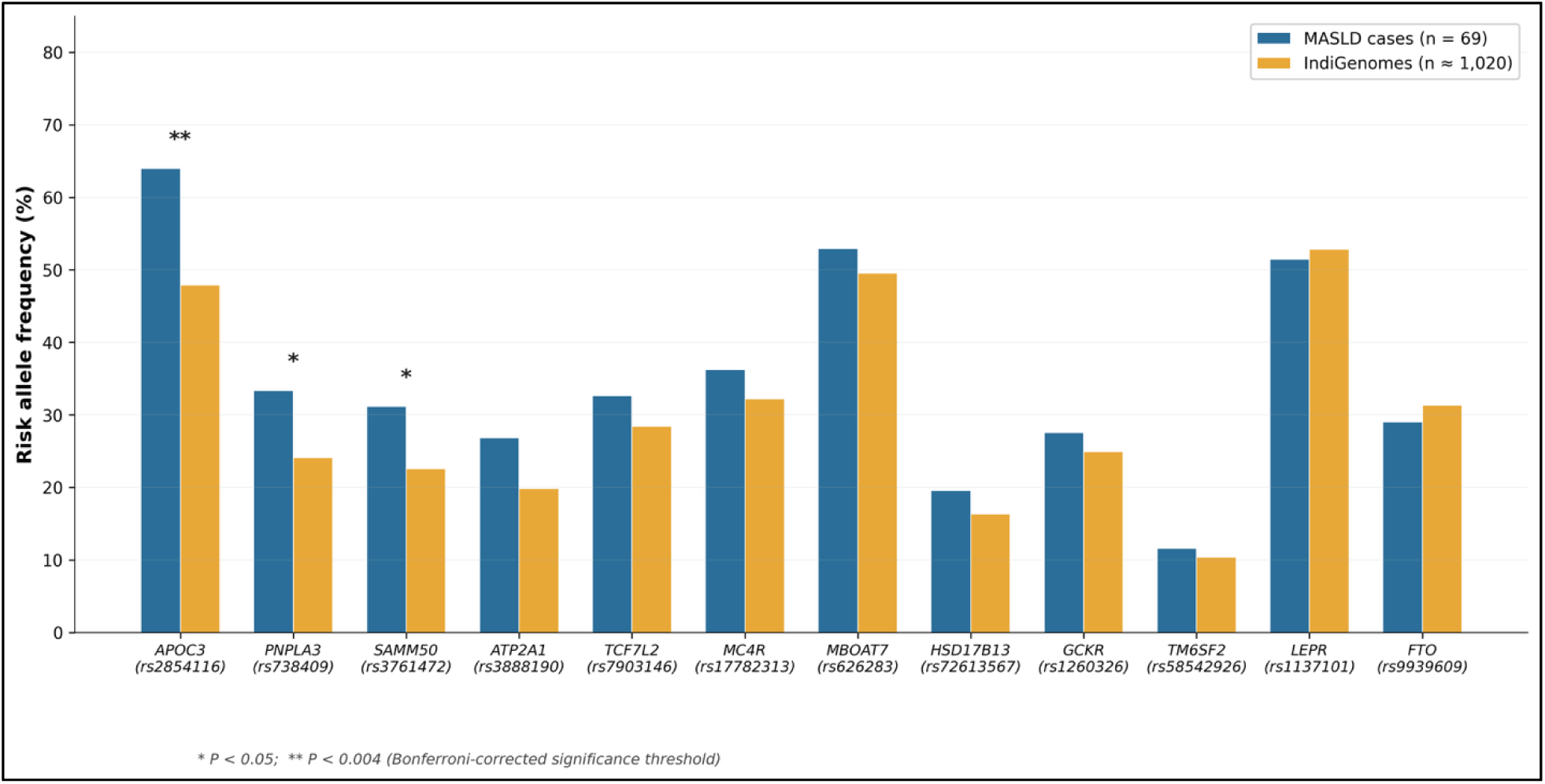
Figure 2. Risk allele frequencies in the MASLD cohort compared with the IndiGenomes Indian reference population. Paired bars show the risk allele frequency (%) for each of the 12 candidate SNPs in MASLD cases (blue, n = 69) and IndiGenomes healthy Indian controls (amber, n ≈ 1,020). SNPs are ordered by decreasing magnitude of case–reference difference. *APOC3* rs2854116 T allele was the only variant significantly enriched after Bonferroni correction (64.0% vs. 47.9%, OR 1.93, P < 0.001). *PNPLA3* rs738409 (33.3% vs. 24.1%, P = 0.019) and *SAMM50* rs3761472 (31.2% vs. 22.6%, P = 0.028) were nominally significant. Allelic odds ratios were calculated using Fisher’s exact test. *P < 0.05; **P < 0.004 (Bonferroni-corrected threshold for 12 comparisons).

**Table 4:**
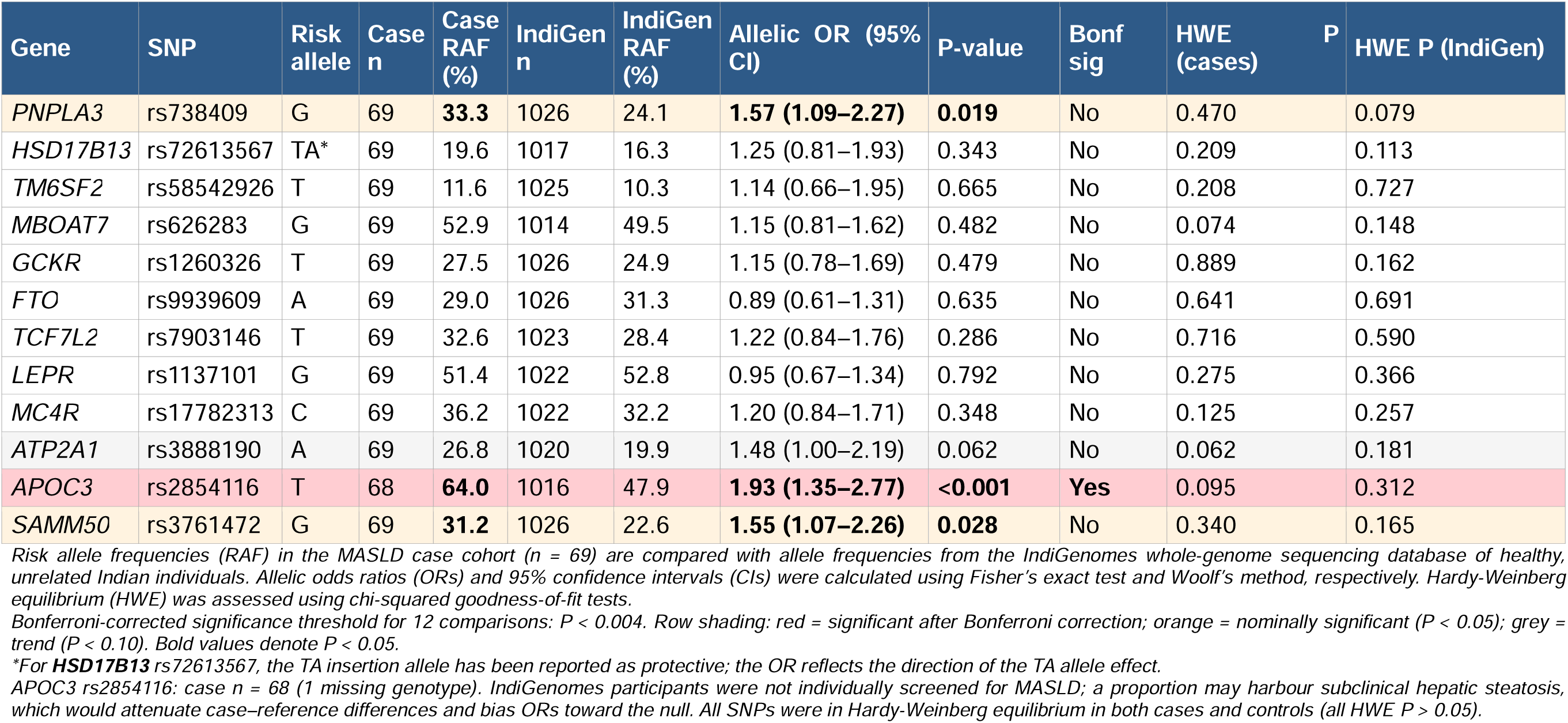
Risk allele frequencies in the MASLD cohort compared with the IndiGenomes Indian reference population.

PNPLA3 rs738409 G allele was nominally enriched in cases (33.3% vs. 24.1%, OR 1.57, 95% CI 1.09–2.27, P = 0.019), as was SAMM50 rs3761472 G allele (31.2% vs. 22.6%, OR 1.55, 95% CI 1.07–2.26, P = 0.028). ATP2A1 rs3888190 showed a borderline trend (26.8% vs. 19.9%, OR 1.48, P = 0.062). The remaining eight SNPs did not differ significantly between cases and IndiGenomes. All SNPs were in Hardy-Weinberg equilibrium in both the case cohort and the IndiGenomes reference, supporting the validity of these comparisons.

### Population-level context: global allele frequency comparison

To place our findings within a broader population-genetic framework, we compared risk allele frequencies across six reference groups: MASLD cases, IndiGenomes, and the five 1000 Genomes super-populations (South Asian, European, East Asian, African, and Admixed American) for all 12 SNPs (Figure 3). The APOC3 rs2854116 T allele was notably higher in our MASLD cohort (64.0%) than in any reference population, including the 1000 Genomes South Asian superpopulation (approximately 44.5%) and IndiGenomes (47.9%), underscoring its marked enrichment in Indian MASLD patients. The PNPLA3 G allele in cases (33.3%) exceeded the IndiGenomes and South Asian reference values (approximately 24–26%) but was substantially lower than in Admixed American (approximately 48%) and East Asian (approximately 40%) populations, consistent with the known ancestry-dependent gradient for this locus. SAMM50 showed a similar pattern of enrichment in Indian MASLD relative to Indian and South Asian references but comparable frequency to East Asian populations. These observations reinforce the necessity of using ancestry-matched controls and caution against extrapolating Western genetic risk models to the Indian context.

**Figure 3:**
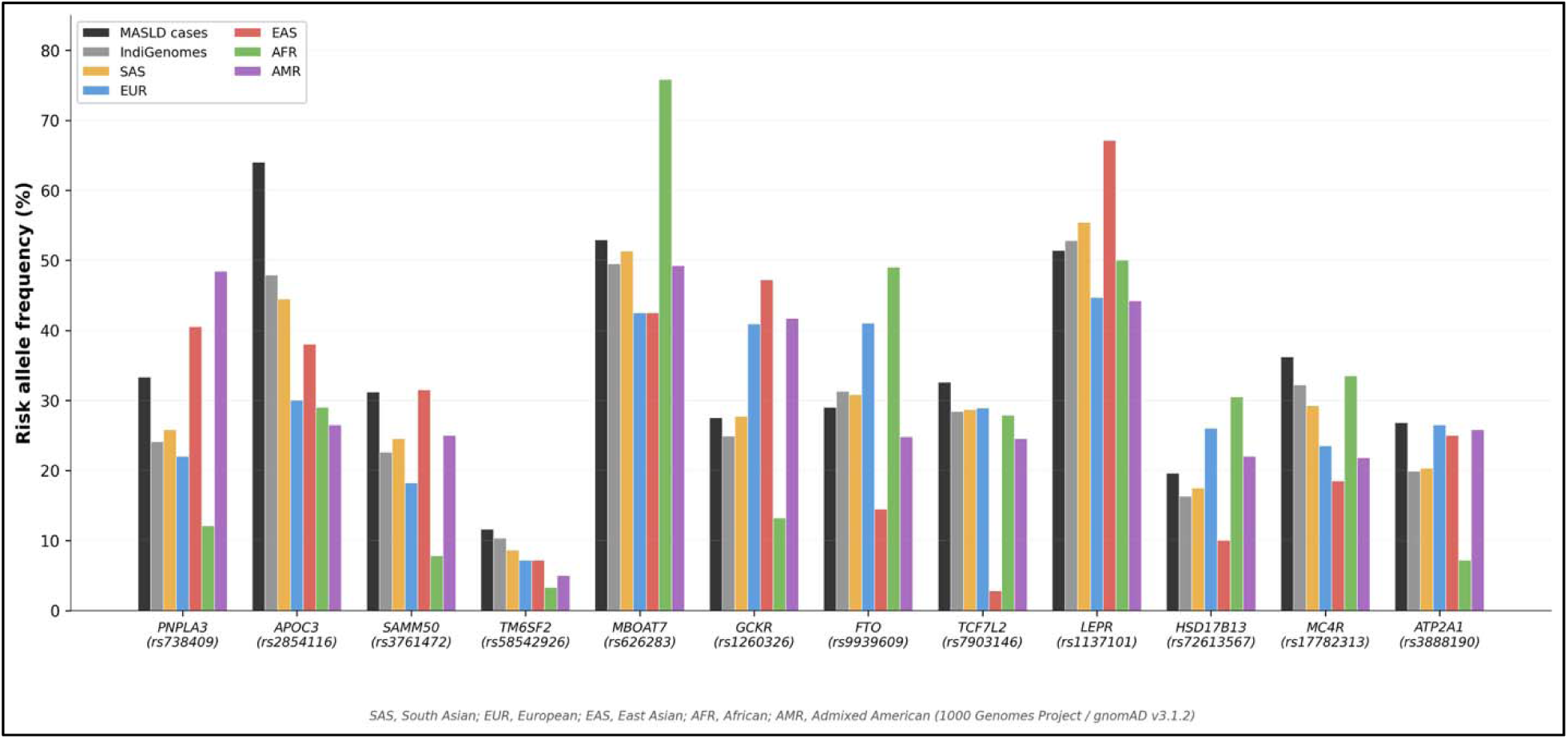
Population-level comparison of risk allele frequencies for 12 candidate SNPs. Risk allele frequencies (%) in the MASLD case cohort, IndiGenomes Indian reference population, and five global super-populations from the 1000 Genomes Project (SAS, South Asian; EUR, European; EAS, East Asian; AFR, African; AMR, Admixed American). The APOC3 rs2854116 T allele was markedly enriched in MASLD cases (64.0%) compared with all reference populations, including IndiGenomes (47.9%) and 1000 Genomes SAS (44.5%). Notable population-specific variation is seen for PNPLA3 (highest in AMR), MBOAT7 (highest in AFR), and LEPR (highest in EAS).

## DISCUSSION

This study presents a systematic evaluation of 12 candidate genetic variants in North Indian patients with MASLD, employing a dual-objective design that separates the genetic determinants of disease severity from those of disease susceptibility. The key finding is a dissociation between these two dimensions: PNPLA3 rs738409 is the dominant predictor of fibrosis within established MASLD, while APOC3 rs2854116 appears to predispose to the disease at the population level without influencing fibrosis progression. This conceptual distinction—between who develops MASLD and who progresses to significant fibrosis—carries implications for both risk stratification and understanding the genetic architecture of the disease in South Asian populations.

The primacy of PNPLA3 rs738409 as a fibrosis-associated variant is consistent with the broader global literature. The I148M substitution impairs hepatocyte triglyceride hydrolysis and promotes lipid droplet accumulation, stellate cell activation, and fibrogenesis through mechanisms that are now well characterised^42,43^. In our cohort, the G allele frequency of 43.4% in the fibrosis group versus 21.0% in the non-fibrosis group yielded an allelic OR of 2.89 (P = 0.006), with a dominant-model OR of 3.94 (P = 0.008). These effect sizes are comparable to those reported in large European and multiethnic meta-analyses^42–44^, and they now extend the evidence to a North Indian MASLD cohort characterised by the metabolic features typical of South Asian patients: high prevalence of central obesity, diabetes, and dyslipidaemia at relatively modest BMI values.

An intriguing observation was the paradoxical inverse relationship between PNPLA3 G carrier status and CAP values (278 vs. 305 dB/m, P = 0.025), which paralleled the same pattern observed in the overall fibrosis-stratified analysis in Table 1. This is consistent with the concept of steatosis regression with fibrosis progression, sometimes termed ‘burnt-out NASH,’ and has been described in the context of PNPLA3-associated disease^45,46^. The finding suggests that in clinical practice, low CAP values should not necessarily be interpreted as benign in patients carrying the PNPLA3 risk allele, as they may reflect advanced fibrotic remodelling rather than absence of hepatic injury.

The SAMM50 rs3761472 and FTO rs9939609 variants showed borderline associations with fibrosis (P = 0.065 and 0.089, respectively) that did not reach statistical significance but are biologically plausible. SAMM50 encodes a mitochondrial outer-membrane protein involved in β-barrel protein assembly and has been implicated in MASLD through its proximity to PNPLA3 on chromosome 22 and through independent functional effects on mitochondrial reactive oxygen species generation^29,47^. The association of SAMM50 G carriers with obesity (97.4% vs. 80.6%, P = 0.040) in our data adds a metabolic dimension to this variant’s phenotypic footprint. FTO’s association with hypertension in our cohort (54.3% vs. 26.5%, P = 0.027) is consistent with its well-established role in adiposity-driven metabolic syndrome^48,49^. These findings, while hypothesis-generating in a cohort of this size, nominate SAMM50 and FTO as candidates for larger replication studies in Indian MASLD.

Perhaps the most important conceptual finding of this study is the behaviour of APOC3 rs2854116. This promoter variant, which upregulates apolipoprotein C-III transcription and thereby inhibits lipoprotein lipase activity, was dramatically enriched in our MASLD cohort compared with IndiGenomes (64.0% vs. 47.9%, OR 1.93, P < 0.001)—the only variant to survive Bonferroni correction in the case–reference comparison. Yet within the cohort, the APOC3 T allele showed no association with fibrosis status (OR 0.96, P = 1.000), LSM, CAP, diabetes, hypertension, or any other measured phenotype. This pattern strongly suggests that APOC3 rs2854116 operates as a susceptibility variant—it predisposes to MASLD development, likely through chronic hypertriglyceridaemia and hepatic lipid influx—but does not govern the subsequent trajectory of fibrosis once the disease is established. This dissociation between susceptibility and severity has precedent in other complex liver diseases^50^ and underscores the importance of distinguishing these two genetic axes in future study designs.

The enrichment of APOC3 rs2854116 in Indian MASLD is particularly relevant to the South Asian metabolic context. Apolipoprotein C-III is a key regulator of triglyceride metabolism, and South Asians are known to carry a disproportionate burden of atherogenic dyslipidaemia characterised by elevated triglycerides, low HDL-cholesterol, and small dense LDL particles^51,52^. The high baseline frequency of the APOC3 T allele in the general Indian population (47.9% in IndiGenomes, versus approximately 30% in European and African populations) suggests that this population already operates at a genetically elevated risk of triglyceride-mediated hepatic steatosis, and that further enrichment of this allele in MASLD patients may represent a threshold effect. This observation aligns with the established efficacy of APOC3 inhibitors (volanesorsen, olezarsen) in reducing hepatic steatosis and triglyceride levels^53,54^, and could inform pharmacogenomic approaches to MASLD management in Indian patients.

Several limitations merit discussion. First, the sample size of 69 patients provides limited statistical power, particularly for variants with modest effect sizes. The Bonferroni-corrected threshold of P < 0.004 was not reached for any within-cohort fibrosis association, including PNPLA3 (P = 0.006). We have reported both nominal and corrected thresholds transparently and characterise significant findings as hypothesis-generating rather than definitive. Second, the IndiGenomes reference population was not screened for MASLD, and a proportion of these ostensibly healthy individuals may harbour subclinical steatosis. This would attenuate case–reference differences and bias odds ratios toward the null, making our observed enrichments conservative estimates. Third, the patients in our cohort were not treatment-naïve—many were on statins, anti-diabetic medications, saroglitazar, vitamin E, or ursodeoxycholic acid—which precluded meaningful analysis of biochemical parameters such as liver enzymes and lipid profiles. We therefore focused on categorical comorbidity diagnoses and physical measurements (FibroScan, anthropometry) that are less susceptible to treatment confounding. Fourth, population stratification within the heterogeneous North Indian population could not be adjusted for in the absence of genome-wide ancestry informative markers. Fifth, environmental factors including dietary composition, physical activity, and microbiome variation were not assessed and may interact with genetic variants to modify disease expression.

Notwithstanding these limitations, this study makes several contributions. It is among the first to systematically separate fibrosis-associated from susceptibility-associated genetic signals in Indian MASLD, providing a conceptual framework for interpreting candidate gene findings. It identifies APOC3 rs2854116 as a population-level MASLD susceptibility marker specific to the Indian genetic context, a finding with potential pharmacogenomic implications. It confirms PNPLA3 rs738409 as a fibrosis predictor in Indian MASLD, consistent with the global literature. And it provides a methodological template for the transparent use of external population references such as IndiGenomes, including explicit acknowledgement of their limitations.

## CONCLUSIONS

In this cohort of 69 North Indian patients with MASLD, we demonstrate a dissociation between genetic determinants of disease susceptibility and fibrosis progression. APOC3 rs2854116 is significantly enriched relative to the Indian general population and likely contributes to MASLD development through triglyceride-mediated hepatic lipid accumulation, while PNPLA3 rs738409 drives fibrosis severity within established disease. SAMM50 rs3761472 shows suggestive dual signals at both levels. These findings highlight the need for ancestry-specific genetic risk profiling in Indian MASLD and provide pilot data to power future multicentre validation studies. Larger prospective cohorts with matched healthy controls and longitudinal follow-up are warranted to confirm these associations, explore gene–gene and gene–environment interactions, and develop clinically actionable polygenic risk scores for the Indian population.

## DATA AVAILABILITY STATEMENT

The genotype and phenotype data supporting the findings of this study are available from the corresponding author upon reasonable request.

## CONFLICT OF INTEREST

Ashish Kumar reports relationships with Roche, Hetero, Viatris, La Renon, Torrent, and Sun Pharma, including receipt of speaking and lecture fees as well as travel reimbursement. These relationships are unrelated to the present study. All other authors declare no conflicts of interest.

## FUNDING

This study did not receive any funding.

## DECLARATION OF GENERATIVE AI AND AI-ASSISTED TECHNOLOGIES IN THE MANUSCRIPT PREPARATION PROCESS

During the preparation of this work the authors used Claude (Anthropic) in order to assist with statistical computation, data visualisation, figure generation, and language editing, and NotebookLM (Google) in order to assist with graphical abstract design. After using these tools, the authors reviewed and edited the content as needed and take full responsibility for the content of the published article.

## Conflict of interest statement

None of the authors declare any conflict of interest.

## Funding information

This work did not receive any funding.

## Credit Authorship statement

The authors contributed to this manuscript as follows:

AK, SC, AA: Conceptualization, Methodology

NT, AK: Original draft preparation

AK: Supervision, Project administration

NT, SC, AA, PS, RV, RH, SJ, RK, TR, MSL, AK: Writing - Review & Editing.

**All authors have read and approved the final version of the manuscript.**

**Genetic susceptibility versus fibrosis progression in North Indian MASLD: distinct roles of APOC3 and PNPLA3 in a candidate gene study**

## ABBREVIATIONS

MASLD: metabolic dysfunction–associated steatotic liver disease;
SNP: single-nucleotide polymorphism;
LSM: liver stiffness measurement;
CAP: controlled attenuation parameter;
OR: odds ratio;
CI: confidence interval;
BMI: body mass index;
HWE: Hardy-Weinberg equilibrium;
T2DM: type 2 diabetes mellitus;
HDL-C: high-density lipoprotein cholesterol;
LDL-C: low-density lipoprotein cholesterol;
IQR: interquartile range;
SD: standard deviation;
GWAS: genome-wide association study;
PCR: polymerase chain reaction;
MASH: metabolic dysfunction–associated steatohepatitis;
RAF: risk allele frequency

## SUPPLEMENTARY MATERIAL

**Supplementary Table S1:**
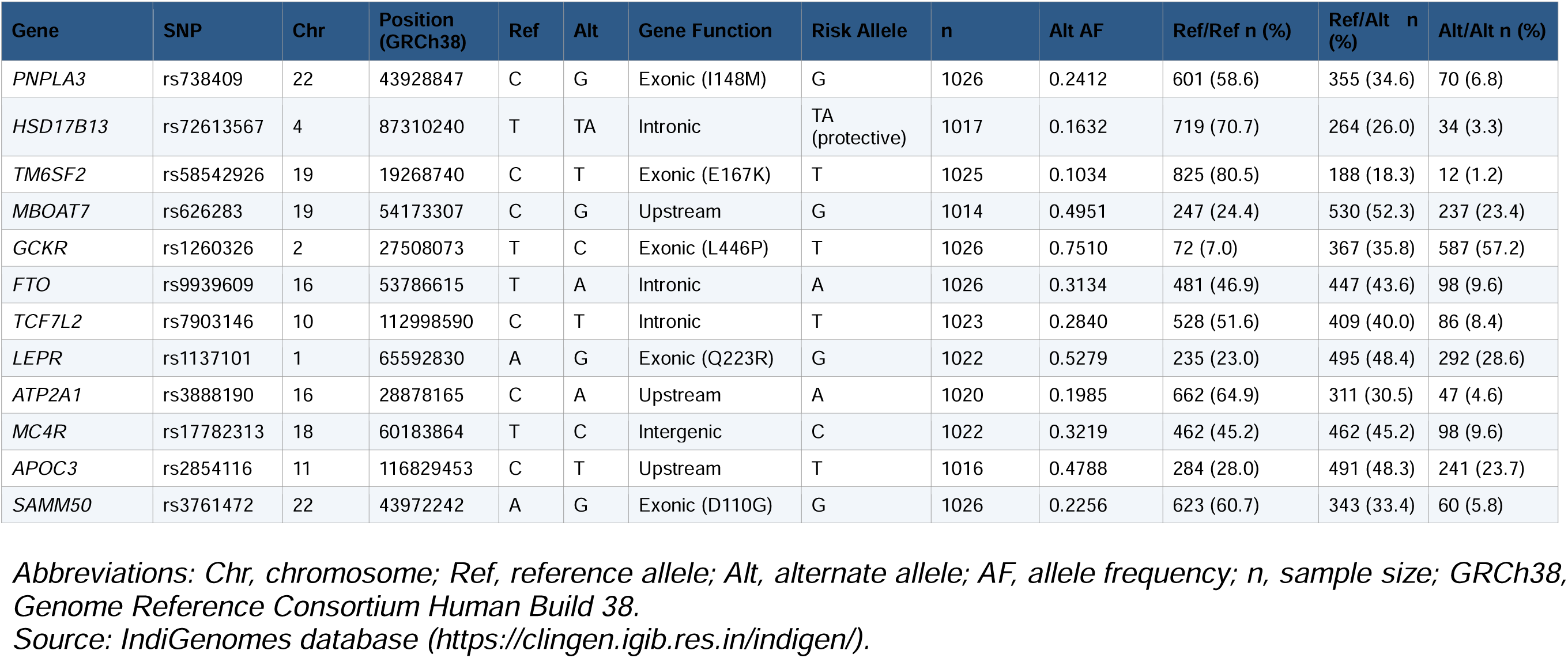
IndiGenomes Reference Population: Allele Frequencies and Genotype Distributions for 12 Candidate SNPs. Allele and genotype frequencies were obtained from the IndiGenomes database, which comprises whole-genome sequencing data from 1,029 healthy, unrelated individuals of diverse Indian ancestry (build GRCh38). Twelve single nucleotide polymorphisms across genes implicated in hepatic lipid metabolism, metabolic regulation, and proteostasis were queried. Alt AF, alternate allele frequency; n, number of individuals with available genotype data for each SNP; Ref, reference allele; Alt, alternate allele. Genotype counts are shown as n (%). Risk alleles are designated based on prior literature and observed direction of effect in the study cohort. For *APOC3* rs2854116, the alternate allele T is the putative risk allele for MASLD; for *HSD17B13* rs72613567, the TA insertion allele has been reported as protective in other populations. Sample sizes vary slightly across SNPs (range: n = 1,014–1,026) depending on data availability in the IndiGenomes database.

## Notes

### Author Declarations

The study protocol was approved by the Institutional Ethics Committee of Sir Ganga Ram Hospital, New Delhi (Approval No. EC/07/25/2735). All participants provided written informed consent prior to saliva collection and genotyping.

